# Burnout Syndrome and coping mechanisms among Prison Officers at Nsawam Medium Security Prison, Eastern Region, Ghana

**DOI:** 10.1101/2025.01.02.25319893

**Authors:** Florence Djoletoe, Philip Teg-Nefaah Tabong

**Affiliations:** Ghana Prisons Service, P. O. Box 129, Accra, Ghana; Department of Social and Behavioural Sciences, School of Public Health, Post Office Box LG 13, College of Health Sciences, University of Ghana, Legon

## Abstract

Workplace burnout negatively affects employees’ productivity, organizational effectiveness, social stability, and personal health and performance. Burnout is a syndrome comprising emotional exhaustion, depersonalization, and loss of personal accomplishment. In Ghana, there are several studies on job-related burnout among health workers but very little on this syndrome in different professional groups like prison officers. Thus, this study assessed burnout syndrome, associated factors, and coping strategies among Prison Officers at Nsawam Medium Security Prison. This was a cross-sectional study involving 266 prison officers from the Nsawam Medium Security Prison. A self-administered questionnaire was used to collect data from participants. Data was collected on sociodemographic characteristics, workplace environment, coping strategies, and burnout. The Maslach Burnout Inventory-Human Services scale was adopted for this study. The data was analysed using STATA 16. Descriptive Statistical methods were used to summarize socio-demographic characteristics of participants. Bivariable analysis using Pearson’s Chi Square was used to determine the association between demographic and work-related factors and burnout. Multivariate logistic regression was used to determine factors associated with burnout at 95% significance level. The prevalence of burnout was 9%. The main causes of burnout perceived by POs were irregular shifts (71.6%), heavy workload (49.0%) and lack of training and opportunities for development (47.7%). Male prison officers had 69% reduction in the odds of burnout compared to their female counterparts (AOR: 0.31; 95% CI= 0.13-0.75, p=0.009). Also, prison officers with diploma/certificate had 89% reduction in odds to burnout compared to those with postgraduate degrees (AOR: 0.11; 95% CI= 0.02-0.69, p=0.019). Talking to colleagues (62.5%), looking forward to off duty (60.1%), talking to family (54.3%), and keeping thoughts and feelings to oneself (42.1%) were the dominant coping strategies. Overall, the prevalence of burnout among prison officers in this study was low. Heavy workloads, erratic shift schedules, and a lack of training and career development chances contribute to prison officers’ burnout. Additionally, shift allocations need to be changed to lessen officer burnout.

## Background

Over the last decades, social changes such as increased job deregulation, job insecurity, conflicting conditions, heavy workload, and work-related stress associated with demands of high standards, quality service, and tight competition [1] have caused modifications at the workplace globally. This resulted in workers becoming emotionally and physically worn out, which has led to an uncontrollable buildup of workload and high levels of burnout. Recognized as a global public health issue, and a problem, before the COVID-19 pandemic, burnout is expected to get far worse with the ongoing crisis [2]. Burnout is a state of physical and emotional tiredness brought on by long-term job-related stress or by working in a physically or emotionally demanding position. According to Maslach and Jackson [3], the syndrome has three components: emotional weariness, depersonalization, and loss of sense of accomplishment. Chronic stress is recognized as the key factor resulting in burnout. In addition, adopting negative coping strategies contribute to the maintenance and worsening of burnout symptoms.

The World Health Organization (WHO) classified burnout as an “occupational phenomenon” in 2019 with a prevalence of 20% in active people [4]. It is reportedly becoming more prevalent among workers worldwide [5]. Geographical regions and professions have been found to have significant differences globally [6]. For instance, it has been found that sub-Saharan African and Asian regions had the greatest prevalence rates of burnout symptoms among healthcare professionals, whereas European and Central Asian regions have the lowest rates [7]. Also, a recent report by Schaufeli, [8] suggests that 10% of the European Union workforce in Europe feels burned out on average. Clearly, the dynamics operating in African prisons are substantially different from those found in Western countries [9]. After almost two decades of ethnographic studies on prisons in East, West, and North Africa, the field has been found to achieve modest advancements in scope and reach as well as relevance and sophistication. Similar to the global picture, there are differences in geographical regions with respect to the inmate population and prison work characteristics resulting in burnout among POs [10,11]. Taken together, under the pressure of overwhelming social, economic, and political challenges observed in most African countries, burnout rates are observed to be high [8].

Burnout may have an impact on an individual’s quality of life [12] as well as the operations of an organization in terms of physical, psychological, behavioral, and defensive symptoms. Prison officers have been recognized as an underserved and at-risk workforce that, in comparison to persons in other professions, has a higher prevalence of mental and physical illness [13]. They are at risk of burnout due to the general adverse conditions characterized by the nature of their work, which includes a variety of institutional and occupational factors [14]. These working conditions increase their chronic exposure to stress, which is the main cause of exhaustion. Coping, the capacity of persons under stress, and the tools at their disposal for dealing with stress are all closely tied to the level of work-related stress.

Currently, burnout is a global public health concern. It was a problem before the COVID-19 pandemic, and because of the current crisis, it is predicted to get far worse [2]. Although as an institution, management of preventive measures would be difficult given the inherent stressors that characterize prison officers’ work, such as irregular work hours, and a heavy workload [15], it is acknowledged that burnout among correctional officers needs urgent attention [14]. Evidence suggests that burnout syndrome and subsequent performance are likely to be impacted by the frequency and length of workdays [16].

Despite indications of research on job-related stress leading to burnout among Ghanaian workers, most of these studies have focused almost exclusively on health professionals. Providing safe custody and the well-being of convicts is the primary duty of the prison officer [17]. Anecdotal evidence show that job requirement puts pressure and strain on the personnel. For example report available at the prison service show that a total of 102 working days were missed in one month for the year 2021[18]. As a result, the affected officers missed an average of 6 days of work during the thirty-day period [18]. This compels the already understaffed, physically fit officers to continue doing the same amount of work, which hinders the Service’s capacity to carry out its mandate. On the other hand, evidence shows that POs experience high levels of burnout because of the distinctive characteristics of their job, far higher than the 46% prevalence rate observed across working populations [19]. About a third (37%) of correctional staff may exhibit burnout related symptoms and this may vary across socio-demographic characteristics and individual coping mechanism [13,16]. In contrast to prisoners in custody, who have several programs to assist them to effectively manage the stress of their living environment, prison officers who spend most of their time in the same space have little resources to help them handle the stress of the prison setting [20]. Moreover, prior research in the Ghanaian prison system mostly ignored the health of the prison officers in favour of the prisoners. This can have negative implications for the Prison Service’s efforts to effectively contribute to the reduction in Ghana’s crime rate [10]. This study therefore investigated the prevalence of burnout, associated factors and coping strategies adopted by POs to provide evidence that may improve health and wellbeing of service officers.

## Methods and Materials

### Ethical Consideration

Approval for the study was sought from Ghana Health Service Ethics Review Committee. Approval was given (GHS-ERC:041/01/23). Ghana Prison Service Administration, Headquarters also gave approval prior to data collection. All participant gave a written consent before data elicitation.

### Study Area

The study was carried out in the Nsawam Medium Security Prison. This Prison is in the Nsawam-Adoagyire Municipality, in the Eastern Region. This area holds the largest prison establishment in Ghana in terms of inmate population and staff strength (GPS and Ghana Prisons Council, 2014). Records show that this prison has 1350 inmates and an estimated staff strength of 608 [21].

### Study Design

A quantitative methods approach was adopted for this study. It was a descriptive cross-sectional study. This type of study involves selecting a cross section of Prison Officers working in the Nsawam Medium Security Prison for data elicitation.

### Study Population

The population for the study comprises Prison Officers of the Ghana Prison Service who perform general duties at the NMSP. Officers actively working at the prison at any point in time may belong to one of these groups; officers in training (Recruit officers or Cadet officers); junior rank (non-commissioned officers) or senior rank (commissioned officers). General duties include escorting, custody, and control of inmates, as well as administrative and supervisory roles (Ghana Prisons Service Ten Year Strategic Development Plan, 2014). POs who have been performing general yard duties for six months or more were recruited.

### Inclusion and Exclusion Criteria

Prison Officers who perform general duties at the NMSP were included in this study. However, those on annual at the time of data collection were excluded.

### Sample Size Determination

Using the Taro Yamane’s formula for known population [22] with an assumption of 95% level of confidence, the population of prison officers as given by the Eastern Regional Command of the Ghana Prisons Service was 608. The Yemane formula for cross-sectional study:

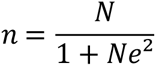

Where n = sample size

N= Total NSMP officer population

e = Margin of error set at 0.05

N= 608 (Ghana Prisons Service, 2020)

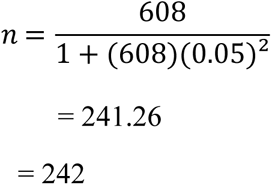

The minimum sample size considering a 10% non-response rate (Nix et al., 2019) was 266.

### Sampling Technique

A proportionate stratified random sampling was adopted in recruiting participants for the study. A list of the officers was obtained from the Eastern Regional Command of GPS and divided into two groups based on their corps (commissioned and non-commissioned respectively). A sample size proportionate to each group is selected using simple random sampling and the result combined to make the sample for the study (Table 1).

**Table 1:**
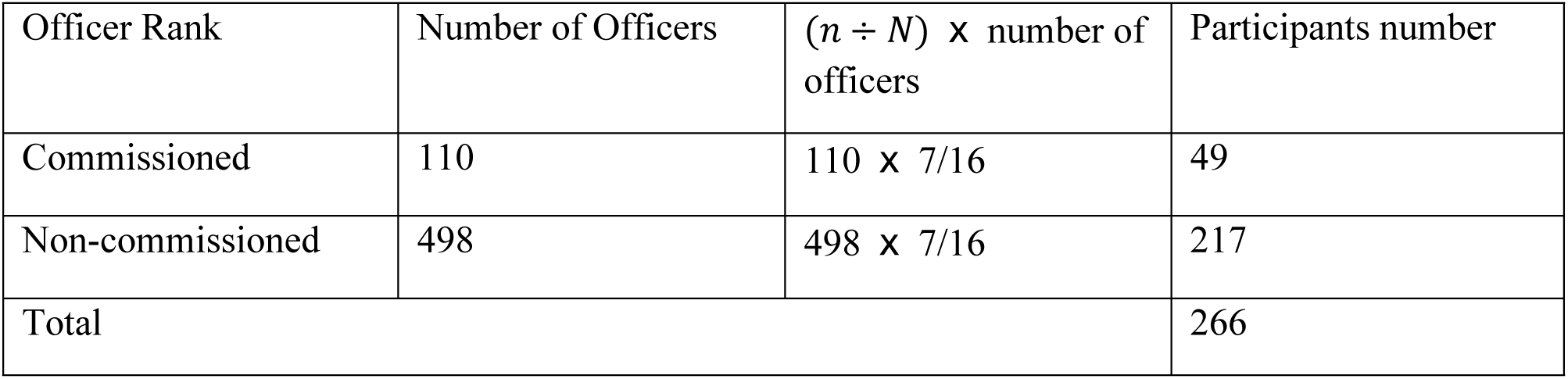
Number of participants required from each corps for the study.

| Officer Rank | Number of Officers | $(n \div N)$ x number of officers | Participants number |
| --- | --- | --- | --- |
| Commissioned | 110 | $110 \times 7/16$ | 49 |
| Non-commissioned | 498 | $498 \times 7/16$ | 217 |
| Total |  |  | 266 |

Number of commissioned officers present = 110

Number of non-commissioned officers present = 498

Total number of officers (n) = 608

Sample size (n) = 267

Sample fraction 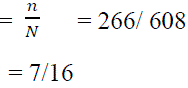

Participants were selected using pieces of paper with the option YES and NO in a bowl from which officers were allowed to pick one. Those who picked yes were given a briefing about the study and participants who consented were recruited for the study. The data collection started on the 5th of February 2023 and completed on the 18th May, 2023.

### Data Collection Tool

Data was collected using self-administered questionnaires assessing burnout among the POs. Both open and closed-ended questions were used to capture information relevant to identifying POs socio-demographics, what they perceive as causes of burnout, and coping mechanisms adopted. The questionnaire was adapted from the Maslach Burnout Inventory-Human Services Survey (MBI-HSS) [23], the Work Environment Scale [24] and the Coping Methods Checklist [25]. These are validated tools and have been used in similar studies with other fields such as health workers in Ghana [26,27].

The questionnaire comprised four sections. The first section included data on socio-demographic factors: age, sex, educational level, marital status, and work characteristics such as: rank, years of service and supervisory role. The second section gathered information on POs’ perception of the workplace using the Work Environment Scale and to identify what officers perceived to be causes of burnout. The Work Environment Scale [24] consists of seventeen statements that measures the social environment of work settings. It assesses employee’s perceptions of workplace using the dimensions of relationships, personal growth, and the system maintenance by either agreeing or disagreeing to the statements.

Burnout was measured using the MBI-HSS that explores the three dimensions of burnout: emotional exhaustion, depersonalization, and professional accomplishment, in the third section.

The adapted 22-item MBI-HSS has each statement rated on a seven-point likert scale that measures how often officers experience a particular feeling from 0 (never) to 6 (everyday). It measures burnout using the three constructs.

EE with 9 items

DP with 5 items

LPA with 8 items

The Coping Methods Checklist was adapted in the last section to gather data on the different interventions adopted by POs to reduce burnout. This adapted seventeen (17) statements tool is useful in assessing perceived efficacy of various coping strategies in relation to chronic job-related stress ([28]. Participants indicated from a list coping strategy which was helpful in managing burnout. However, “others” option was available for people to indicate coping strategies that were not part of the list.

### Data Collection Technique

After one of the prison facility’s weekly meetings, participants were given an information sheet prior to the administration of the questionnaire. This was a summary of the study in simplified language.

Participants were allowed questions for clarification of doubts which were answered appropriately. Consenting participants were made to complete a hardcopy version of the questionnaire.

### Data Analysis

Of the 266 participants, 245 questionnaires were returned and after data cleaning, 243 had been filled and used for analysis. Data were exported to STATA version 16 and coded for analysis (StataCorp, 2019).

Descriptive statistical measures were used to describe the socio-demographic factors and work characteristics of participants. The scores in each dimension are used to characterize the level of burnout as high, moderate, or low.

The MBI-HSS tool, a 7-point Likert scale, was used to measure the three constructs of burnout.

0 = Never

1 = At least a few times a year

2 = At least once a month

3 = Several times a month

4 = Once a week

5 = Several times a week

6 = Everyday

Scores for each construct were then computed and categorized. For EE, the scale was categorized as Low (≤16), Moderate (17-26) and High (≥27). Additionally, the DP scale was categorized as Low (≤6), Moderate (7-12) and High (≥13). Finally, PA was also categorized as Low (<31), Moderate (32-38) and High (≥39) (Table 2).

**Table 2.** Source used for burnout levels assessment.

|  | Low | Moderate | High |
| --- | --- | --- | --- |
| Emotional exhaustion | $\leq 16$ | 17-26 | $\geq 27$ |
| Depersonalization | ≤6 | 7-12 | ≥ 13 |
| Personal accomplishment | ≥ 37 | 31-36 | ≤30 |

Scores for EE, DP and PA were summarized as means and standard deviations. Burnout was computed as a composite variable using scores from participants. The definition of burnout adopted for the study as provided by Abdo et al., [29] and Dubale et al., [30] was high scores on EE and DP and low scores on PA. Thus, participants with high EE, high DP and low PA were said to be burned out while those who did not fall in this categorization were classified as not burned out.

Additionally, assessing data on perceived causes of burnout and coping skills adopted by POs using the Coping Methods Checklist were also presented as frequencies and percentages.

Chi-square/Fisher’s Exact where appropriate were used to determine the relationship between socio-demographic characteristics, EE, DP, PA and Burnout. P-values less than 0.05 were considered statistically significant. Multivariate logistic regression was used to determine factors associated with burnout. Based on this, a dichotomous variable was created “0” (no burn out) and “1” (burn out). The outcome of interest was burnout. Strengths of association between independent variables and burnout were determined using crude odds ratio. Variables with p-value of <0.05 in unadjusted logistic regression model (COR) were considered for inclusion into the multivariate logistic regression analyses, adjusted logistic regression (AOR). Issues relating to cofounders were addressed using multilinear collinearity using the Variance of Inflation Factor (VIF) command in Stata. To test for goodness of fit of Model II, the likelihood ratio test was used to examine the likelihood of data under the full model as against the likelihood of the data under a model with reduced number of independent variables. A p-value for the overall model obtained was less than 0.05. Thus, it was concluded the model was good. P-values less than 0.05 were considered statistically significant. The results have been presented in tables and graphs.

## Results

### Socio-demographic and work characteristics of prison officers

The total number of prison officers recruited for this study was two hundred and sixty-six (266). Of this, 245 questionnaires were returned and two hundred and forty-three (243) used for analysis (Table 3). The participation rate of this study was 92%.

**Table 3:** Sociodemographic characteristics of prison officers.

| Characteristic | Frequency | Percentage |
| --- | --- | --- |
| Age | Mean (SD)= 32.4 (7.5) |  |
| 20-30 | 115 | 47.29 |
| 31-40 | 102 | 42.00 |
| 40+ | 26 | 10.71 |
| Sex |  |  |
| Female | 77 | 31.69 |
| Male | 166 | 68.31 |
| Marital status |  |  |
| Single | 140 | 57.61 |
| Married | 98 | 40.33 |
| Divorced/widowed | 5 | 2.05 |
| Education |  |  |
| SHS | 133 | 54.73 |
| Diploma/certificate | 45 | 18.52 |
| Degree | 53 | 21.81 |
| Postgraduate | 12 | 4.94 |
| Work experience |  |  |
| <5 years | 148 | 60.90 |
| 5-10 years | 39 | 16.00 |
| 10+ years | 56 | 23.10 |
| Rank |  |  |
| Junior | 190 | 78.19 |
| Senior | 53 | 21.81 |
| Schedule Head |  |  |
| No | 196 | 80.66 |
| Yes | 47 | 19.34 |
| Shift Allocation |  |  |
| Early | 60 | 24.69 |
| Late | 16 | 6.58 |
| Main | 144 | 59.26 |
| Night | 23 | 9.47 |
| Have children |  |  |
| No | 126 | 51.85 |
| Yes | 117 | 48.15 |
| Have chronic condition |  |  |
| No | 221 | 90.95 |
| Yes | 22 | 9.05 |

The study revealed that while about half (47.29%) of the respondents were between the ages 20-30 years, their ages range between eighteen to fifty-eight (18-58) years. The mean age of officers was 32.40 ± 7.55 years. Majority (68.31%) were male. Additionally, more than half were single (57.61%), had attained senior high school education (54.73%), have been working in prison service for less than five years (60.90%) and were junior ranked officers (78.19%). Moreover, 19.34% worked as schedule heads. The predominant shift (59.26%) engaged by officers within the past 6 months was the main shift (8am-4pm). Less than half (48.15%) indicated they had children and a greater proportion reported not having chronic conditions (90.95%).

### Prevalence of burnout among prison officers

The three dimensions of burnout were measured using the 22-item Maslach Burnout Inventory–Human Services Survey tool (MBI-HSS). The summary of scores for each construct is summarized in Table 4. Overall, 25.9% of the officers had low emotional exhaustion (EE), 42.0% had high emotional exhaustion. Furthermore, the distribution of the three categories of depersonalization (DP) were similar for the officers: Low (35.4%), Moderate (32.1%) and High (32.5%). Regarding personal accomplishment, the majority (68.7%) of the participants had low personal accomplishment while only 14% had high personal accomplishment (Figure 1).

**Figure 1:**
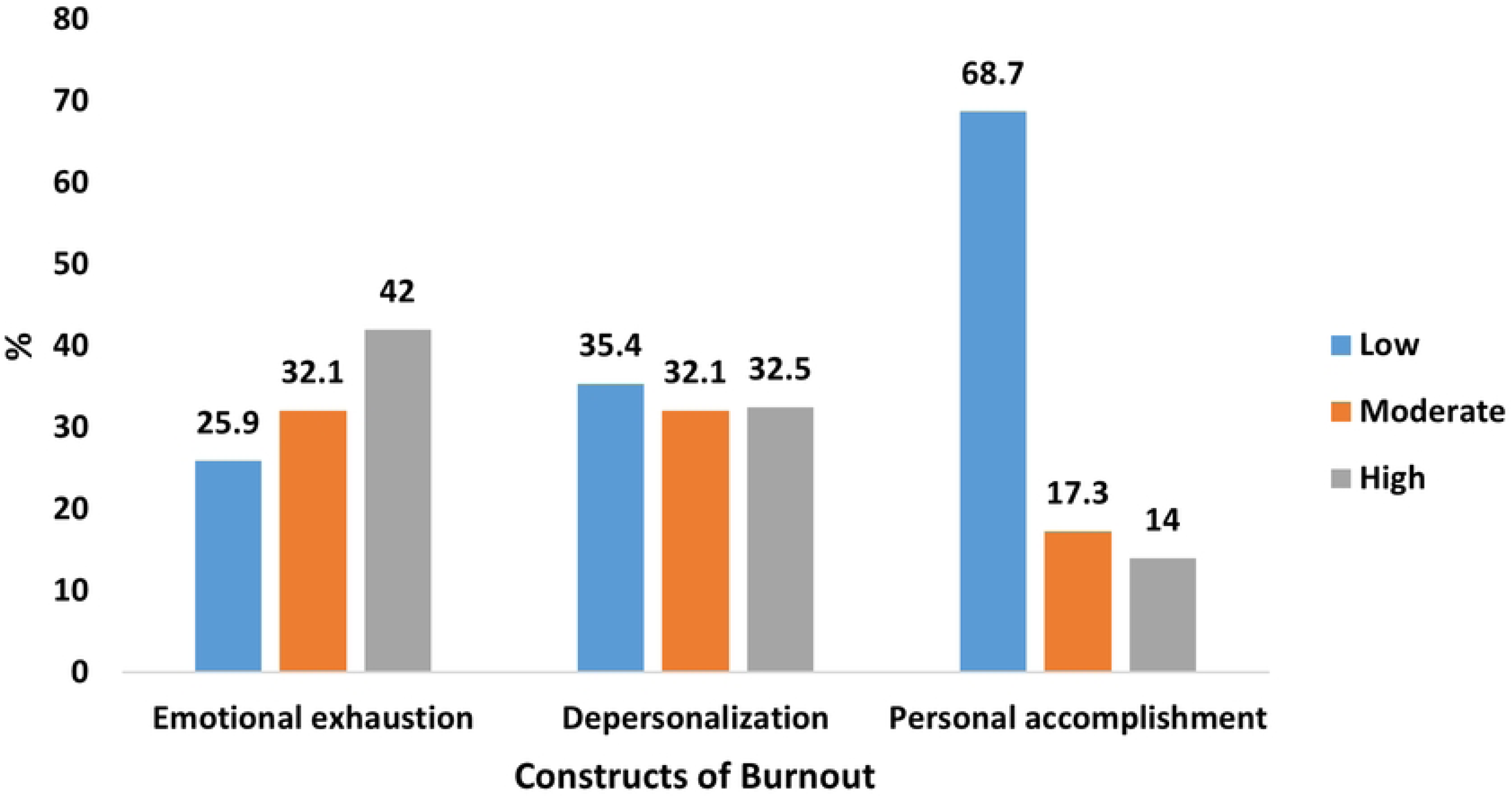
Distribution of Dimensions of Burnout among Prison Officers.

**Table 4:** scores of dimensions of burnout among prison officers.

| Variable | Minimum | Maximum | Mean | SD |
| --- | --- | --- | --- | --- |
| Emotional Exhaustion | 0 | 54 | 24.9 | 12.5 |
| Depersonalization | 0 | 30 | 10.6 | 7.4 |
| Personal Accomplishment | 0 | 48 | 26.3 | 10.7 |

Burnout was defined as a high EE (≥ 27) or high DP (≥ 10) and low PA (≤33). Overall, the prevalence of burnout syndrome among the prison officers was 9% (Figure 2).

**Figure 2.**
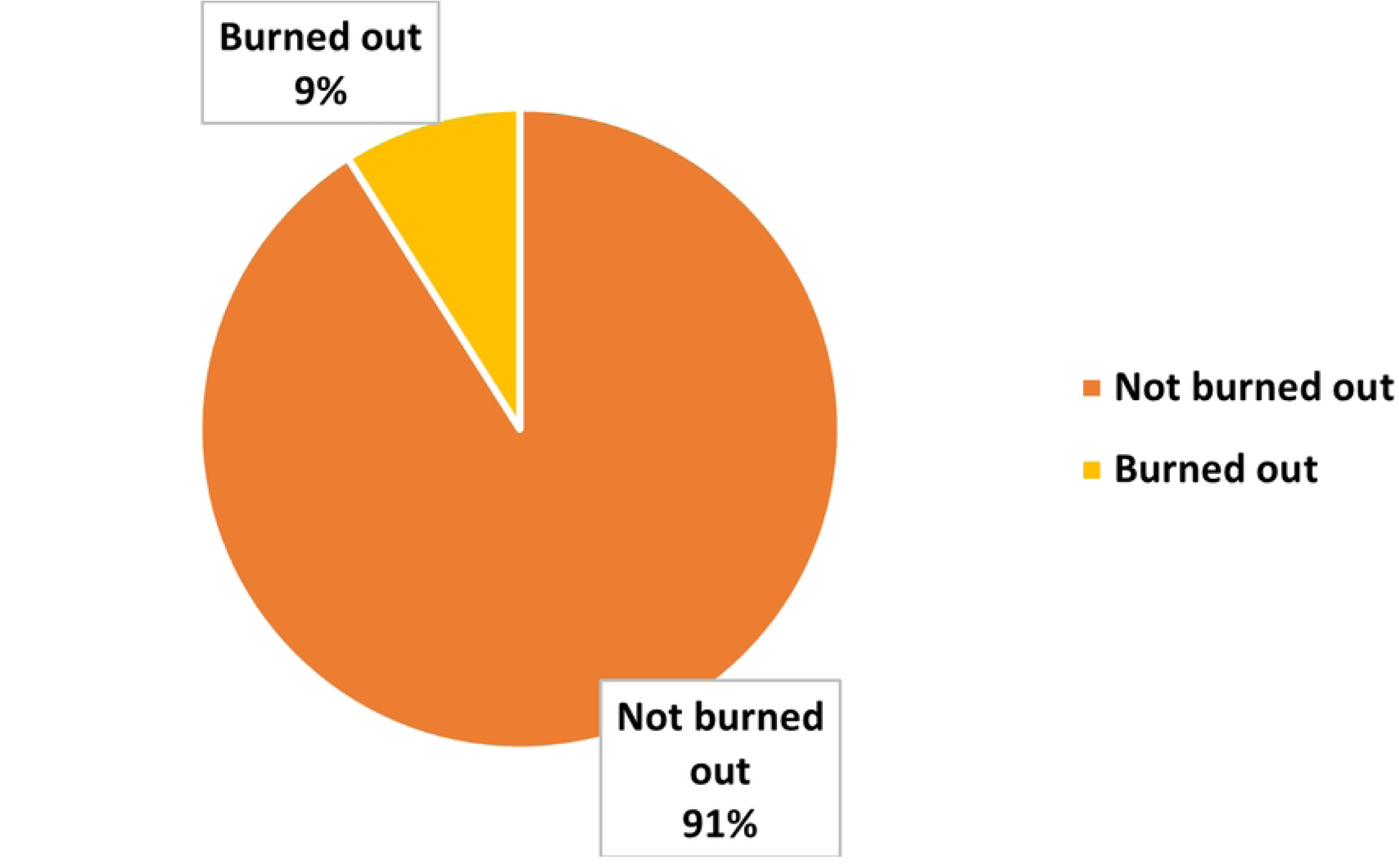
Prevalence of Burnout among Prison Officers.

### Perceived causes of burnout among prison officers

Perceived causes of burnout were assessed among POs in this study (Figure 3). The respondents identified various causes they perceived which include irregular shifts (71.6%), heavy workload (49.0%) and lack of training and opportunities for development (47.7%). Others included lack of autonomy (35.8%), no variety and poor social status (28.0%).

**Figure 3.**
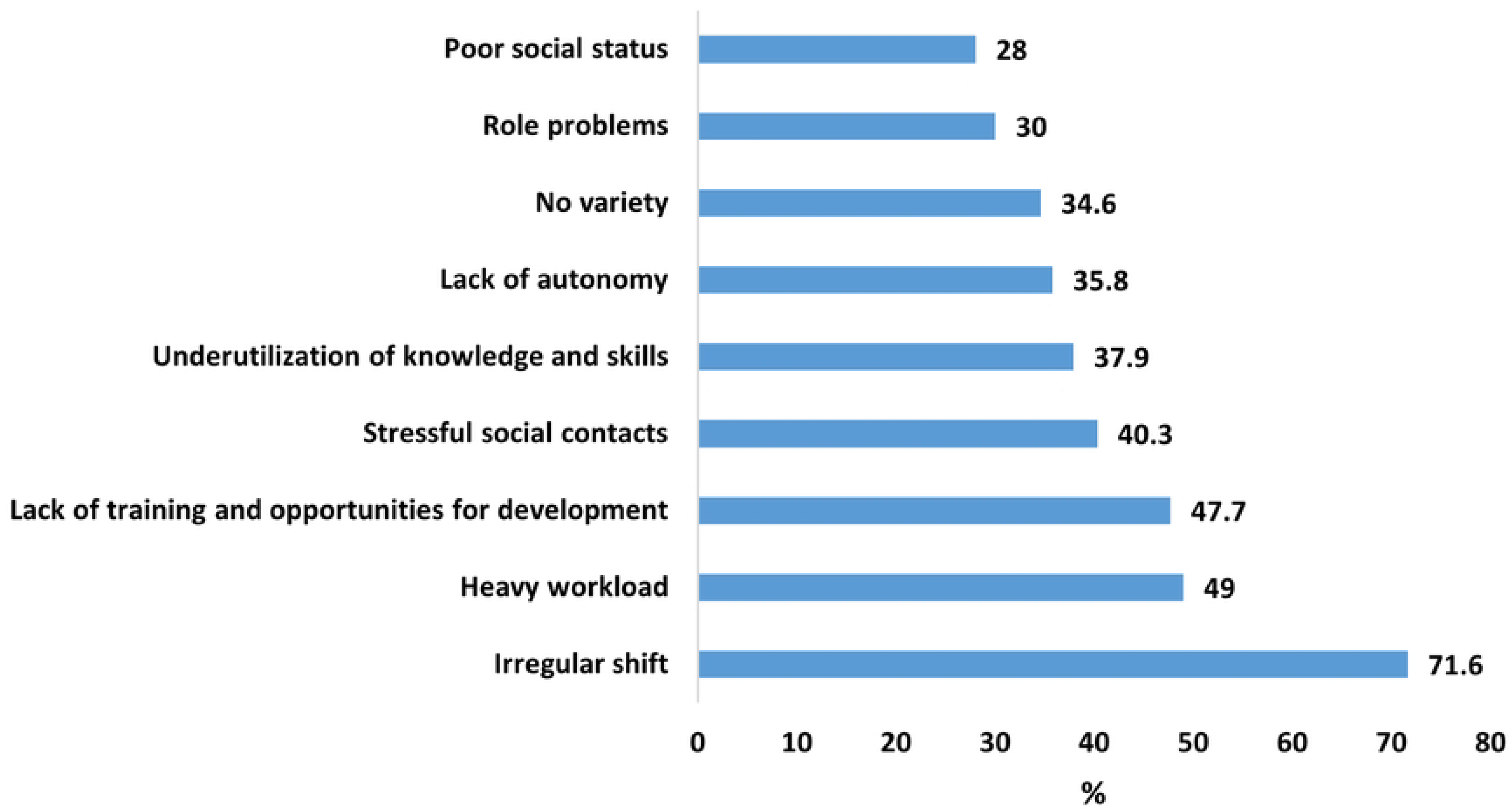
Perceived Causes of Burnout among prison officers.

### Assessment of prison work environment

Table 5 represents how POs rate the prison work environment. More than half (56.4%) disagreed that there is a positive and supportive culture and emotional climate, agreed they felt like part of a team (59.3%) and felt challenged when assigned tests to stretch their efforts (58.4%). Additionally, 59.7% disagreed that their efforts were recognized and received constructive feedback (55.1%). Furthermore, 74.1% agreed they took pride in their work and workplace, felt they were in control of their work (71.2%) and felt accepted and valued by their colleagues (72.0%). On the contrary 50.6% disagreed they trusted their superiors to be there for them, disagreed they felt safe sharing their plans with the officer in charge and disagreed the rewards for success were greater than the penalties (62.1%).

**Table 5.** Rating of prison work environment.

| Statement | Agree<br>N(%) | Disagree<br>N(%) |
| --- | --- | --- |
| The culture and emotional climate of the prison is generally positive and supportive ie welfare, health, | 106(43.6) | 137(56.4) |
| I feel like I am part of a team (shared vision, goals, efforts) | 144(59.3) | 99(40.7) |
| I feel challenged and assigned allocations that inspire, test and stretch my efforts | 142(58.4) | 101(41.6) |
| My efforts are recognized in a tangible way e. g. awards | 98(40.3) | 145(59.7) |
| I receive constructive feedback in a way that emphasizes positives rather than negatives | 109(44.9) | 134(55.1) |
| Innovation is expected of me, and I am encouraged to take initiative | 127(52.3) | 116(47.7) |
| I have clear-cut and non-contradictory instructions on my job description | 123(50.6) | 120(49.4) |
| I believe in and take pride in my work and my workplace | 180(74.1) | 63(25.9) |
| I feel in control of my work and capable of competently carrying out my duty duties | 173(71.2) | 70(28.8) |
| I feel accepted and valued by my colleague officers | 175(72.0) | 68(28.0) |
| I feel accepted and I'm treated with courtesy, listened to and invited to express my thoughts and feelings by my superiors. | 135(55.6) | 108(44.4) |
| The station's administrative team provides an environment in which I feel safe and secure | 115(47.7) | 128(52.7) |
| I believe administration considers my needs and preference when making decisions. | 99(40.7) | 144(59.3) |
| My professional judgement is respected by my superiors; I have adequate freedom to exercise my judgement and expertise | 94(38.7) | 149(61.3) |
| I trust my superiors to be there for me and back me up. | 120(49.4) | 123(50.6) |
| I feel safe sharing my plans, programs and policies with my OIC | 118(48.8) | 124(51.2) |
| The rewards for success are greater than the penalties for failure | 92(37.9) | 151(62.1) |

### Association between dimensions of burnout and demographic characteristics

The Pearson’s chi-square test was used to assess the association between demographic characteristics of respondents and the three constructs of burnout syndrome (Table 6). Results showed that all other socio-demographics, aside shift allocation was significantly associated with emotional exhaustion (p=0.010). The percentage of emotional exhaustion was significantly higher among POs who run main shift (68.0%) compared to those with early (22.5%), night (17.7%) and late (5.9%) shifts. Furthermore, officer rank was also significantly associated with depersonalization (p=0010). The percentage of depersonalization was significantly higher among junior officers (76.0%) as compared to senior officers (24.0%). No sociodemographic characteristic had a statistically significant association with personal accomplishment.

**Table 6.** Association between constructs of burnout and socio-demographic characteristics.

|  | Emotional Exhaustion |  |  |  | Depersonalization |  |  |  | Personal Accomplishment |  |  |  |
| --- | --- | --- | --- | --- | --- | --- | --- | --- | --- | --- | --- | --- |
|  | Low<br>N(%) | Moderate<br>N(%) | High<br>N(%) | p-value | Low<br>N(%) | Moderate<br>N(%) | High<br>N(%) | p-value | Low<br>N(%) | Moderate<br>N(%) | High<br>N(%) | p-value |
| Age |  |  |  |  |  |  |  |  |  |  |  |  |
| 20-30 | 30(47.6) | 33(42.3) | 52(51.0) | 0.657 | 42(48.8) | 29(37.2) | 44(55.7) | 0.154 | 80(47.9) | 19(45.2) | 16(47.1) | 0.170 |
| 31-40 | 26(41.3) | 38(48.7) | 38(37.2) |  | 33(38.4) | 41(52.6) | 28(35.4) |  | 73(43.7) | 14(33.3) | 15(44.1) |  |
| 40+ | 7(11.1) | 7(9.0) | 12(11.8) |  | 11(12.8) | 8(10.3) | 7(8.9) |  | 14(8.4) | 9(21.4) | 3(8.8) |  |
| Sex |  |  |  |  |  |  |  |  |  |  |  |  |
| Female | 16(25.4) | 22(28.2) | 39(38.2) | 0.165 | 27(31.4) | 21(26.9) | 29(36.7) | 0.419 | 58(34.7) | 10(23.8) | 9(26.5) | 0.309 |
| Male | 47(74.4) | 56(71.9) | 63(61.8) |  | 59(68.6) | 57(73.1) | 50(63.3) |  | 109(65.3) | 32(76.2) | 25(73.5) |  |
| Marital status |  |  |  |  |  |  |  |  |  |  |  |  |
| Single | 30(47.6) | 1(1.3) | 2(2.0) | 0.440 | 46(53.5) | 41(52.6) | 53(67.1) | 0.061 | 96(57.5) | 25(59.5) | 19(55.9) | 0.989 |
| Married | 31(49.2) | 29(37.2) | 38(37.2) |  | 36(41.9) | 37(47.4) | 25(31.6) |  | 68(40.7) | 16(38.1) | 14(41.2) |  |
| Divorced/widowed | 2(3.2) | 48(61.5) | 62(60.8) |  | 4(4.6) | 0(0.0) | 1(1.3) |  | 3(1.8) | 1(2.4) | 1(2.9) |  |
| Education |  |  |  |  |  |  |  |  |  |  |  |  |
| SHS | 32(50.8) | 42(53.8) | 59(57.8) | 0.928 | 42(48.8) | 45(57.7) | 46(58.2) | 0.265 | 95(56.9) | 21(50.0) | 17(50.0) | 0.453 |
| Diploma/certificate | 12(19.0) | 16(20.5) | 17(16.7) |  | 14(16.3) | 15(19.2) | 16(20.5) |  | 31(18.6) | 8(19.1) | 6(17.7) |  |
| Degree | 16(25.4) | 15(19.2) | 22(21.6) |  | 27(31.4) | 14(17.9) | 12(15.2) |  | 31(18.6) | 13(30.9) | 9(26.5) |  |
| Postgraduate | 3(4.8) | 5(6.4) | 4(3.9) |  | 3(3.5) | 4(5.1) | 5(6.3) |  |  |  |  |  |
| Work experience |  |  |  |  |  |  |  |  |  |  |  |  |
| <5 years | 42(66.7) | 47(60.3) | 59(57.8) | 0.196 | 56(65.1) | 42(53.9) | 50(63.3) | 0.073 | 103(61.7) | 24(57.1) | 21(61.8) | 0.770 |
| 5-10 years | 9(14.3) | 17(21.8) | 13(12.7) |  | 8(9.3) | 20(25.6) | 11(13.9) |  | 29(17.4) | 6(14.3) | 4(11.8) |  |
| 10+ years | 12(19.0) | 14(17.9) | 30(29.4) |  | 22(25.6) | 16(20.5) | 18(22.8) |  | 35(20.9) | 12(28.6) | 9(26.5) |  |
| Rank |  |  |  |  |  |  |  |  |  |  |  |  |
| Senior | 21(33.3) | 20(25.6) | 25(24.5) | 0.435 | 33(38.4) | 14(17.9) | 19(24.0) | 0.010* | 42(25.1) | 14(33.3) | 10(29.4) | 0.539 |
| Junior | 42(66.7) | 58(74.4) | 77(75.5) |  | 53(61.6) | 64(82.1) | 60(76.0) |  | 125(74.9) | 28(66.7) | 24(70.6) |  |
| Schedule Head |  |  |  | 0.308 |  |  |  | 0.085 |  |  |  | 0.466 |
| No | 47(74.6) | 63(80.8) | 86(84.3) |  | 65(75.6) | 61(78.2) | 70(88.6) |  | 137(82.0) | 31(73.8) | 28(82.3) |  |
| Yes | 16(25.4) | 15(19.2) | 16(15.7) |  | 21(24.4) | 17(21.8) | 9(11.4) |  | 30(18.0) | 11(26.2) | 6(17.7) |  |
| Shift Allocation |  |  |  |  |  |  |  |  |  |  |  |  |
| Early | 21(33.3) | 16(20.5) | 23(22.5) | 0.010* | 22(25.6) | 20(25.6) | 18(22.8) | 0.220 | 42(25.2) | 12(28.6) | 6(17.6) | 0.380 |
| Late | 4(6.4) | 6(7.7) | 6(5.9) |  | 5(5.8) | 4(5.1) | 7(8.9) |  | 13(7.8) | 1(2.4) | 2(5.9) |  |
| Main | 36(57.1) | 53(68.0) | 55(53.9) |  | 54(62.8) | 49(62.8) | 41(51.9) |  | 100(59.9) | 22(52.4) | 22(64.7) |  |
| Night | 2(3.2) | 3(3.8) | 18(17.7) |  | 5(5.8) | 5(6.4) | 13(16.5) |  | 12(7.2) | 7(16.7) | 4(11.8) |  |
| Have children |  |  |  |  |  |  |  |  |  |  |  |  |
| No | 27(42.9) | 41(52.6) | 58(56.9) | 0.214 | 43(40.0) | 35(44.9) | 48(60.8) | 0.125 | 89(53.3) | 19(45.2) | 18(52.9) | 0.640 |
| Yes | 36(57.1) | 47(47.4) | 44(43.1) |  | 43(50.0) | 43(55.1) | 31(39.2) |  | 78(46.7) | 23(54.7) | 16(47.1) |  |
| Chronic condition |  |  |  |  |  |  |  |  |  |  |  |  |
| No | 59(93.6) | 71(91.0) | 91(89.2) | 0.628 | 77(89.5) | 73(93.6) | 71(89.9) | 0.612 | 155(92.8) | 37(88.1) | 29(85.3) | 0.295 |
| Yes | 4(6.4) | 7(9.0) | 11(10.8) |  | 9(10.5) | 5(6.4) | 8(10.1) |  | 12(7.2) | 5(11.9) | 5(14.7) |  |

\*Statistically significant at $p < 0.05$

### Factors associated with burnout among prison officers

A Pearson chi square test showed sex was significantly associated with burn out (p=0.016). The percentage of burnout was significantly higher among female officers (54.5%) compared to their male counterparts (45.5%). A logistic regression model was used to predict socio-demographic factors associated with burnout (Table 7). The model predicted sex and education to be significant factors associated with burnout among the prison officers. Male prison officers had 69% reduction in the odds of burnout compared to their female counterparts (AOR: 0.31; 95% CI= 0.13-0.75, p=0.009). Also, prison officers with diploma/certificate had 89% reduction in the odds of burnout compared to those with postgraduate degrees (AOR: 0.11; 95% CI= 0.02-0.69, p=0.019).

**Table 7.**
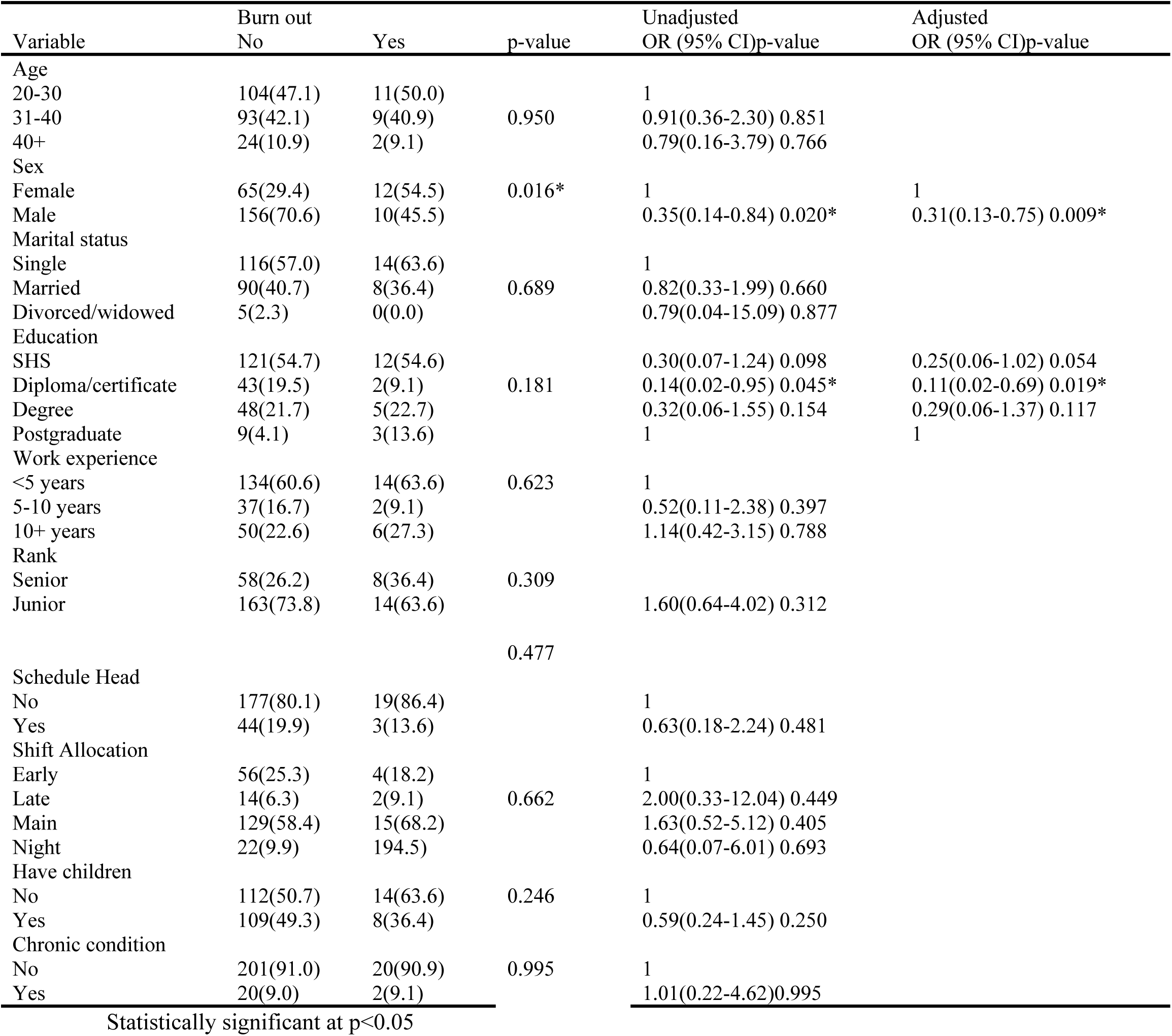
Factors associated with burnout among prison officers.

### Coping strategies for dealing with stress among prison officers

Participants in this study outlined various coping mechanisms for dealing with stress (Figure 4). The main coping strategy was talking to colleagues (62.5%). Others reported looking forward to off duty (60.1%), talking to family (54.3%) and keeping thoughts and feelings to oneself (42.1%). Finally, alcohol use (29.2%) was the least coping strategy reported by respondents.

**Figure 4.**
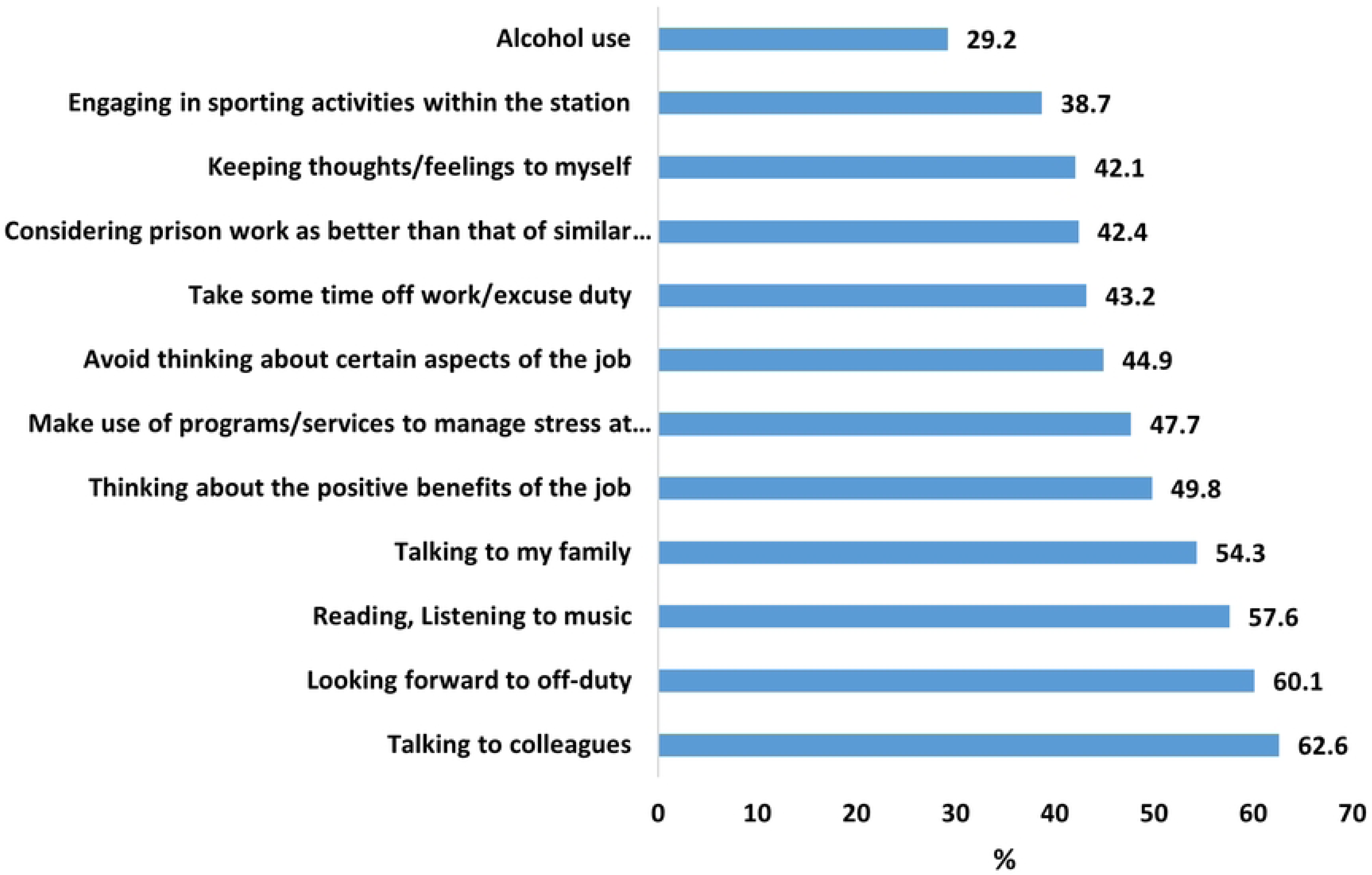
Coping Strategies for Dealing with Burnout Among Prison Officers.

## Discussion

### Prevalence of Burnout Among Prison Officers

The overall prevalence of burnout syndrome among prison officers involved in this study was found to be 9%. This is lower than the over 30% prevalence rate reported by some studies [31,32]. It is consistent with findings indicative of low to moderate levels of burnout among POs [12], confirming that all POs will experience burnout in varying proportions [14]. Though conducted among different worker groups, this study findings are similar to one by Odonkor and Frimpong in 2020 [33], among Ghanaian health workers where burnout level was reported to be low. Also, Dodam et al., [27] in the year 2022 found that in comparison to Ghanaian clinicians, non-clinicians experienced burnout syndrome at a lower rate of 15.64%. This generally low burnout prevalence could be partly due to cultural contexts like in Ghana where indirect communication is favored, and emotions are internalized as opposed to people in Western settings where explicit communication is the norm. Respondents may thus limit the expression of emotions and psychological states.

Furthermore, although more than half (56.4%) of the participants disagreed that a positive and supportive culture and emotional climate existed at the workplace, a good number of them (59.3%) felt a part of the team and could draw from the social support indirectly to reduce stress leading to burnout hence the low level [34]. Most officers in this study reported low levels of personal accomplishment and depersonalization, but high scores of emotional exhaustions). There is a difference in prison systems between African and Western countries [9] and this disparities in the findings can be attributed to the difference in geographical locations and characteristics of individuals in different locations [11,35]. In addition, prison officers involved in the current study reported some level of satisfaction and pride in their work, hence the low levels of depersonalization identified.

Regardless of the evidence to show that in resource-constrained settings such as Ghana and Ghanaian prison setting, POs are likely to show more signs of burnout resulting in lack of motivation and dedication lowering organizational commitment, unproductive attitudes and counterproductive behaviours like encouraging inmates to commit crimes while they are in prison custody which will negatively impact the reformative and rehabilitative process, [10,36], this study found that in addition the low burnout observed among POs, the pride and satisfaction that majority of them reported would result in stronger institutional commitment given the right environment. This implies the need for strengthening self-care issues among professional groups prone to burnout through improvement in the health and the psychosocial environment to prevent negative work outcomes such as illnesses, absenteeism, and mental illnesses in future.

### Perceived Causes of Burnout among Prison Officers and Work Characteristics

Among respondents of this study, the majority perceived irregular shifts as the cause of chronic stress/burnout. This was followed by a heavy workload and lack of training and opportunities for development. The least perceptible source of stress was poor social status. These findings were consistent with findings of a review of about 140 published documents that reported that stressors of prison officers include their work demands, low levels of professional skills, and lack of organizational structures in prisons [37]. A study by Misis et al.,[38] also indicated that a key source of stress among prison officers included characteristics of inmates, which is related to prison officers’ workload. Just as the majority of studies attribute correctional officer burnout to the prison environment as against personal characteristics [39,40], findings from this study showed that institutional sources of stress regarding work schedules and the organizational climate were perceived more than those linked to psycho-social factors and logistical support.

In assessing the prison work environment, most prison officers (56.4%) reported negatively to a positive and supportive culture and emotional climate. This was evidenced in the number of officers who indicated that their efforts were not recognized and failed to receive constructive feedback from their superiors. In addition, officers did not trust their superiors to be there for them and did not feel safe sharing their plans with their superiors/officer in charge. Consequently, the responses from prison officers who partook in this study indicated that the penalties meted out far exceeded the rewards given for successful work done. Boateng and Hsieh [41], in their study on burnout among POs in some Ghanaian prisons found that lack of supervisors’ support contributes to high-stress levels among officers. The findings of their study are inconsistent with the findings of this study as participants were noted to have responded positively about their supervisors. In another study conducted to evaluate the impact of the work environment on prison staff, the role of supervisors was emphasized as being a cardinal factor to increase or decrease stress among correctional officers [34]. These can be attributed to the fact that supervisors encourage the officers not to give up in line of duty.

It is recognized that the inherent stressful nature of prisons coupled with resource constraint make it difficult for GPS administration to manage burnout preventive measures. However, the differences in how POs perceive conditions within the prison, as a result of difference in shifts assigned, duty points and objective assessment of conditions experienced [42,43] call for improvement in working conditions, modification of workloads among others [44].

In addition to supervisory influence, peer influence also affects stress and burnout experienced by correctional officers. Prison officers in this research agreed to the feeling of being part of a team. Consequently, majority reported taking pride in their work and their workplace, felt accepted and valued by their colleagues, and felt in control of their work. In a study to explore the relationship between social support and job burnout among correctional staff, family and friends’ support was found to significantly reduce burnout among correctional staff [34]. These findings can be attributed to the fact that social support provides a sense of belonging and a feeling of being cared for by one’s social circle. This is also confirmed by the commonest coping style adopted by officers in this study. These findings suggest the importance of designing and implementing programs focused on identification of specific aspects of work schedules, work environment and strategies to address them.

### Influence of Socio-demographic factors on Burnout

Depersonalization among prison officers was found to be significantly associated with officers’ rank. According to the findings, junior officers were more likely to experience depersonalization compared to senior officers. This finding can be attributed to the fact that junior officers are just getting started on the job and may experience some form of repulsion from the inmates. On the MBI-HSS, depersonalization describes being emotionally detached from clients as a means of dealing with emotional stressors [45], which is a strategy junior officers are likely to employ at the workplace.

Furthermore, supervisory roles are usually held by senior officers and corroborate findings that show that supervisors perceive less job-related stress and more job satisfaction than their colleagues who were not in authority [34]. Also, consistent with the findings of this study, Morgan et al., [46] reported that depersonalization among prison officers decreases with increasing age.

### Burnout Coping Mechanism

Evidence suggests that although organizational and individual-based tools are available for correctional officers to manage stress, Maricuţoiu et al., [47] revealed that 96% of the interventions adopted were individualized for each worker, using cognitive-behavioral interventions, interpersonal skills interventions, relaxation interventions, and other strategies. The commonest coping strategy prison officers utilized in dealing with stress in this study was found to be talking to colleagues.

Possible reasons for this are that POs’ shift system characterized by long hours allow them to engage with colleagues and prisoners frequently, and the fact that the majority of respondents in this study did not find social interactions in the yard stressful corroborates this. Evidence shows that burnout is more likely to occur when interpersonal relationships are poor [48] and the contrary low burnout rate found in this study could be attributed to the good interpersonal relations between officers and officers and inmates.

Other notable coping strategies included looking talking to relatives, looking forward to days off and taking advantage of employee assistance programs. Most researchers report that prison officers cope well with stress through support from relatives, friends, and colleagues. This line of discussion is confirmed by the results of this study which is reflected in the top three coping strategies adopted by participants. It can be explained by the fact that although the mechanism of coping adopted by the PO is influenced by purely personal factors, it may also be favored by the work context [49]. In addition, some forms of social support have been found to be advantageous to certain constructs of burnout syndrome [34]. All these individualized approaches; cognitive-behavioral interventions and interpersonal skills interventions.

The organizational strategies employed by respondents include engaging in sporting activities, off-duties and employee assistance programs. Alcohol use was identified as the least coping strategy utilized by prison officers

### Socio-demographic Factors and Burnout

In determining factors associated with burnout among prison officers, sex was found to have a significant association in this study. Female officers were more likely to experience burnout compared to their male counterparts. In this regard, this study showed similar trend with a study among correctional officers in Spain [12] and United States [50]. Results from the current study showed that majority of the respondents were males. It is expected that in a male prison like NMSP where the proportion of male officers is higher than female officers and a significant proportion of the workload and prison duties is shouldered by them [18], they are more likely to experience burnout. However, this inconsistent finding could be attributed to the fact that men in Africa have reportedly been shown to be more prone to mental health issues because of internalizing masculine ideals. They engage in constrictive emotionality through defensive strategies by downplaying their mental health issues and status because of ingrained and institutionalized views [51]. Perhaps this may have reflected in their responses in this study. This phenomenon is also characterized in their health seeking behaviours [52]. Furthermore, research mentions how female officers may find the prison environment more stressful given the hazardous and risky nature of prison environment [53]. In their study, Ghaziri et al., [50] found that female correctional nurses reported higher levels of verbal abuse and sexual harassment by prisoners than male nurses in the same prison.

Of particular interest is how female officers at NSMP who are assigned duty points that do not put them in direct contact with inmates for long relative to their male counterparts such as escorting inmates and block duties and are restricted to the office-space (administrative duties) and workshops are more burned out [21]. An explanation for this observation is that those assigned duty points at the offices and workshops run the main shift (8am–4pm) which could be associated with higher workload; they have deal with the poor logistical support of administration and are more prone to the perceived influence from administrative supervisors [8] unlike their male colleagues. Also, just like female prisoners who constitute a minority among inmate population whose needs may be neglected, there is the likelihood that these female officers may have unmet needs predisposing them to burnout.

### Conclusions

The study underscores the need to pay attention to irregular shift, heavy workload, and regular training and opportunities for development to reduce burnout syndrome among POs. The study further concludes POs experience high levels of emotional exhaustion and depersonalization and low levels of personal accomplishment. In addition, emotional exhaustion was common among Officers running night shifts whilst Junior Officers experienced more depersonalization. POs adopted positive coping mechanisms.

### Authors’ Contributions

FD conceived the idea, wrote the proposal, participated data collection and analysis. PT-NT provided technical support on the methodology, data analysis and wrote the draft manuscript. All authors read and approved the final version of the manuscript.

## Data Availability

The data is available upon request made to the Administrator of Ghana Health Service Ethics Review Committee

## Acknowledgements

We acknowledge the support of Prison Service for allowing us to conduct this study.

